# The spatial distribution of coupling between tau and neurodegeneration in amyloid-β positive mild cognitive impairment

**DOI:** 10.1101/2023.04.13.23288533

**Authors:** Belfin Robinson, Shankar Bhamidi, Eran Dayan, the Alzheimer’s Disease Neuroimaging Initiative

**Affiliations:** Biomedical Research Imaging Center, University of North Carolina at Chapel Hill, Chapel Hill, North Carolina 27514, USA; Department of Statistics and Operations Research, University of North Carolina at Chapel Hill, Chapel Hill, North Carolina 27599-3260, USA; Department of Radiology, University of North Carolina at Chapel Hill, Chapel Hill, North Carolina 27599, USA

**Keywords:** amyloid-β, tau, Alzheimer’s disease, multilayer networks, Braak

## Abstract

Synergies between amyloid-β (Aβ), tau, and neurodegeneration persist along the Alzheimer’s disease (AD) continuum. This study aimed to evaluate the extent of spatial coupling between tau and neurodegeneration (atrophy) and its relation to Aβ positivity in mild cognitive impairment (MCI). Data from 409 subjects were included (95 cognitively normal controls, 158 Aβ positive (Aβ+) MCI, and 156 Aβ negative (Aβ-) MCI) Florbetapir PET, Flortaucipir PET, and structural MRI were used as biomarkers for Aβ, tau and atrophy, respectively. Individual correlation matrices for tau load and atrophy were used to layer a multilayer network, with separate layers for tau and atrophy. A measure of coupling between corresponding regions of interest/nodes in the tau and atrophy layers was computed, as a function of Aβ positivity. The extent to which tau-atrophy coupling mediated associations between Aβ burden and cognitive decline was also evaluated. Heightened coupling between tau and atrophy in Aβ+ MCI was found primarily in the entorhinal and hippocampal regions (i.e., in regions corresponding to Braak stages I/II), and to a lesser extent in limbic and neocortical regions (i.e., corresponding to later Braak stages). Coupling strengths in the right middle temporal and inferior temporal gyri mediated the association between Aβ burden and cognition in this sample. Higher coupling between tau and atrophy in Aβ+ MCI is primarily evident in regions corresponding to early Braak stages and relates to overall cognitive decline. Coupling in neocortical regions is more restricted in MCI.

## INTRODUCTION

Alzheimer’s disease (AD), the most common form of neurodegeneration, has become a key contemporary public health concern (Nichols et al., 2019). While the cause of this disease is still unknown, it is believed to develop from the accumulation of the extracellular amyloid-β (Aβ) peptide and from tangles of hyperphosphorylated tau, which lead to synaptic impairment, neuronal loss (atrophy), and consequently to cognitive and behavioral decline (Kumar et al., 2015). The leading model as to how these pathological processes bind together is known as the amyloid cascade hypothesis (Ricciarelli and Fedele, 2017). According to this influential framework, Aβ pathology initiates alterations in tau which then lead to neurodegeneration and to the cognitive and behavioral manifestations of AD (Karran et al., 2011).

The serial and linear structure of the amyloid cascade hypothesis has, nevertheless, been challenged in the literature. In particular, studies suggest that Aβ, tau, and neurodegeneration (atrophy) could have synergistic effects in AD pathogenesis (Busche and Hyman, 2020). Yet, the extent of spatial coupling between alterations in AD pathological biomarkers, specifically in biomarkers for tau and atrophy remains uncertain(LaPoint et al., 2017; Mak et al., 2018; Sepulcre et al., 2016; Xia et al., 2017). On the one hand, studies have reported large degrees of spatial overlap throughout the brain between tau burden, as assessed using positron emission tomography (PET), and magnetic resonance imaging (MRI)-based measures of atrophy in both cognitively normal controls and individuals with AD (Xia et al., 2017). On the other hand, studies have found more restricted spatial coupling between tau and atrophy, which may emerge from heterogeneity in patterns of tau spread (Mohanty et al., 2023). Moreover, the majority of studies that examined interactions between tau and atrophy were either in normal controls or in individuals with AD (Digma et al., 2019; Liu et al., 2021). The extent of coupling between these biomarkers in individuals with MCI and AD pathologic changes, who can be considered as being at the prodromal stages of AD(Jack et al., 2018), remains less understood.

Over the last decade, there has been significant interest and methodology development in the study of network valued data over the same node set (e.g., regions in the brain), but across multiple layers (De Domenico, 2017; Kivelä et al., 2014). These methods may help in clarifying the extent of coupling that exists between tau and atrophy, as they offer additional insight on complex relationships within and between variables in multiple layers, which may be missed by studies looking at single-layer covariance. Multilayer networks allow to model and study complex heterogeneous relationships between entities within a system and variation of these relationships across layers (Boccaletti et al., 2014; Kivelä et al., 2014). In the context of brain networks, multilayer network models were used for understanding the relationships between brain structure, function, and dynamics across multiple scales, both in healthy brain and in AD (Cai et al., 2020; De Domenico, 2017; Guillon et al., 2019). We reasoned that the extent of spatial coupling between tau and atrophy could be modeled using multilayer networks, since this approach can allow to inspect interactions both within and between network layers.

In the current study, we used a cross-sectional sample of participants with MCI (n=314), as well as data from cognitively normal (CN) participants (n=95) to reconstruct single-subject multilayer networks that represent tau and atrophy as separate layers, and the interactions among these two biomarkers in-between layers. More specifically, tau PET and structural MRI (atrophy) data from 72 regions of interest (ROIs) were first extracted from MCI and CN participants **(Figure 1A)**. Tau and atrophy data were then z-score transformed relative to the means and standard deviations from the entire pool of CN participants **(Figure 1B)**. Subsequently, individual-subject covariance matrices were computed for each participant **(Figure 1C)** and used to reconstruct tau and atrophy networks for each participant after minimally thresholding the edge weights to retain all positive weights in the networks **(Figure 1D)**. The tau and atrophy networks were then modeled as multilayer networks and grouped according to Aβ positivity **(Figure 1E)**. This allowed us to study the interaction between the tau and atrophy layers **(Figure 1F)** at the presence and absence of Aβ positivity. We further examined whether the extent of coupling between tau and atrophy differed among transentorhinal, limbic and isocortical ROIs, defined based on the Braak staging system (Braak et al., 2011). These steps allowed us to evaluate our method for assessment of coupling against an established staging scheme for tau pathology.

**Figure 1:**
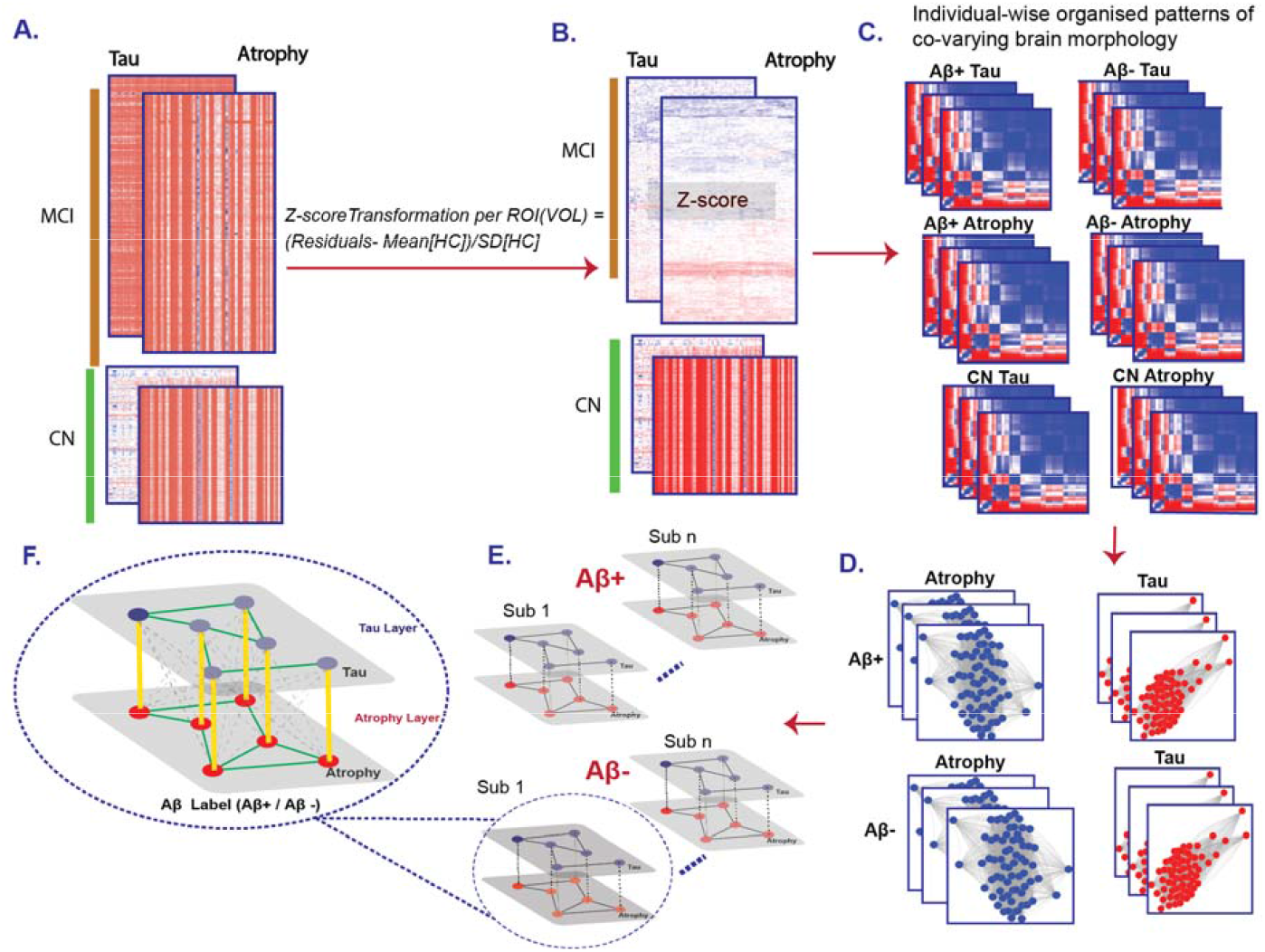
Modeling framework. (A.) Participant’ (MCI, CN) tau standardized uptake value ratios (SUVRs) were calculated in 72 regions of interest (ROIs) (B.) Volumetric measures from the same 72 ROIs were used as measures of atrophy. Data was Z-score transformed using means and standard-deviations (SD) from controls (Value – Mean [CN]/ SD[CN]) (C.) Individual-subject structural covariance matrices were reconstructed for tau and atrophy (D.) Covariance matrices for tau and atrophy were then modeled as single layer networks for each individual participant. (E.) Multilayer networks with tau and atrophy serving as single layers were reconstructed and labeled according to Aβ positivity. (F.) Inter-layer (yellow), Intra-layer (green) and cross-layer (grey-dotted) edges in a multilayer network. MCI – Mild Cognitive Impairment, CN – cognitively normal controls.

## METHODS AND MATERIALS

### Participants

Data used in this study were obtained from the Alzheimer’s Disease Neuroimaging Initiative (ADNI) dataset (https://ida.loni.usc.edu), including the ADNI-1, ADNI-GO, and ADNI-2 cohorts. A total of 314 MCI participants who had ^18^F-florbetapir and ^18^F-flortaucipir PET data were included in the study **(Table 1)**. Data from 95 CN participants was additionally used to aid in the reconstruction of individual-subject graphs/networks for each of the MCI participants. ADNI’s native inclusion and exclusion criteria were used in both the CN and MCI groups. Participants with MCI were further divided into Aβ positive (Aβ+) and negative (Aβ-), based on an established cutoff **(SUVR > 1.11)**, computed relative to the whole cerebellum reference region (Landau et al., 2012b; S.M. et al., 2013). All CN participants were amyloid and tau negative. All subjects provided written informed consent and the procedures were all approved by the local Intuitional Review Boards.

**Table 1.** Characteristics of data used in the study.

| Measure | Aβ + | Aβ - | CN |
| --- | --- | --- | --- |
| Age (years) * | 71.13 ± 6.96* | 70.28 ± 6.71* | 70.45 ± 5.75 |
| Sex | Female: 81 (51)<br>Male: 77 (49) | Female: 72 (46)<br>Male: 84 (54) | Female: 51 (54)<br>Male: 44 (46) |
| Education (years) | 16.34 ± 2.60 | 16.52 ± 2.65 | 16.84 ± 2.56 |

Values are given as mean ± standard deviation or percentages (%); \* denotes significant group difference (p<0.05); Aβ - amyloid-β

### Imaging data analysis

Regional florbetapir PET summary data were obtained from ADNI as derived variables (Landau et al., 2013, 2012a). In short, T1 weighted native-space images were processed with FreeSurfer v7.1.1, and a cortical summary region was defined for each subject, based on frontal, anterior and posterior cingulate, lateral parietal, and lateral temporal ROIs. The T1 images were coregistred to florbetapir PET scans, which allowed to extract PET data from cortical ROI. Standardized uptake value ratio (SUVR’s) were then calculated for the cortical summary region, based on the whole cerebellum reference region. SUVR values from the cortical summary region were then used to define Aβ burden and positivity. Regional summary data based on flortaucipir PET were also obtained from ADNI as derived variables. In short, MPRAGE images were parcellated into a set of 72 ROIs (Braak et al., 2011, 2006; Braak and Braak, 1995) **(See Supplementary Figure 1A, B)** using FreeSurfer v7.1.1 based on the Desikan-Killiany protocol (Desikan et al., 2006). Flortaucipir images were co-registered to the corresponding MPRAGE images to determine the mean regional flortaucipir uptake within each ROI. SUVRs for each of 72 ROIs were then calculated, by dividing uptake values by a cerebellar reference region. Finally, grey matter volumes extracted from the same 72 ROIs using FreeSurfer v7.1.1 were used as measures of regional atrophy.

### Network reconstruction

Regional tau uptake and grey matter volume data from MCI subjects were considered for analysis (**Figure 1A**). Data from CN subjects were further used to aid in the reconstruction of single-subject networks/graphs for each subject with MCI (**Figure 1A**). In this procedure, individual covariance networks in the target group, are reconstructed based on their deviation from an averaged network based on a group of controls (Yun et al., 2020). First, an averaged covariance network was reconstructed from a group of CN subjects (n=95), separately for tau and atrophy. Atrophy and tau data from each MCI subject were then normalized via a z-score transformation using the mean and standard deviation of the averaged, CN-based, tau and atrophy networks (**Figure 1B**). This allowed for the reconstruction of single covariance matrices (Yun et al., 2020, 2015) for each MCI subject (**Figure 1C**). The covariance matrix is a nROI × nROI matrix, where for each index [x,y] we compare the z-scores of the corresponding tau and atrophy ROIs. The equation is given below:

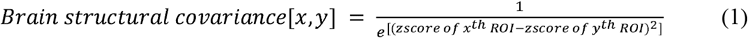

Higher values in ROIs within these matrices denote high covariance compared to the corresponding ROIs from the averaged CN-based networks. The matrices generated for tau and atrophy were structured as single-layer networks/graphs **(Figure 1D)**. The single-layer graphs were then layered into a two-layered graph with tau and atrophy as separate layers **(Figure 1E)**. This form of representation allowed us to examine and compare both intra-layer (green colored edges in **Figure 1F**), and inter-layer edges (yellow-colored edges in **Figure 1F**). While the former type of edges corresponds to covariance for ROIs in the tau and atrophy networks separately, the latter type of edges, which connect the nodes across layers, allow to examine interactions between tau and atrophy. Moreover, unlike multiplex networks, where inter-layer edges connect the same nodes across layers, the multilayer network representation used here also incorporates inter-layer edges connecting across nodes (grey dotted edges in **Figure 1F**). The multilayer networks were generated using R (Version 4.2.1) (Team, 2021), with the igraph (Version: 1.3.5) and muxviz (Version: 3.1) (de Domenico et al., 2015) libraries.

### Interlayer coupling score

A key objective in the current study was to assess the extent of regional/spatial coupling between tau and atrophy in the presence and absence of Aβ positivity. To that effect we have computed a coupling score between the tau and atrophy layers, based on the distance between the layers (Shimada et al., 2016). As a measure of distance, we used Euclidean distance, as it was previously used to assess coupling between network node sets (Liu et al., 2022). First, the partitioned distance between the tau and atrophy layers was calculated. The resulting distance matrix D was used to calculate the coupling score. The distance between identical ROIs across layers was defined as (D_r_), whereas interlayer edges, connecting different ROIs across the layers were defined as (D_b_). The coupling score was computed to measure the relative coupling between tau and atrophy, as the ratio between a spatially coupled edge and all other non-coupled edges:

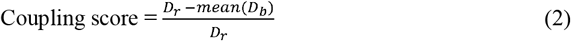

Partitioned Euclidian distance was computed using the pdist (Version 1.2.1) library (https://github.com/jeffwong/pdist) in R (Version 4.2.1). A toy example of the procedure for calculating coupling scores is illustrated in **supplementary Figure 2**.

**Figure 2:**
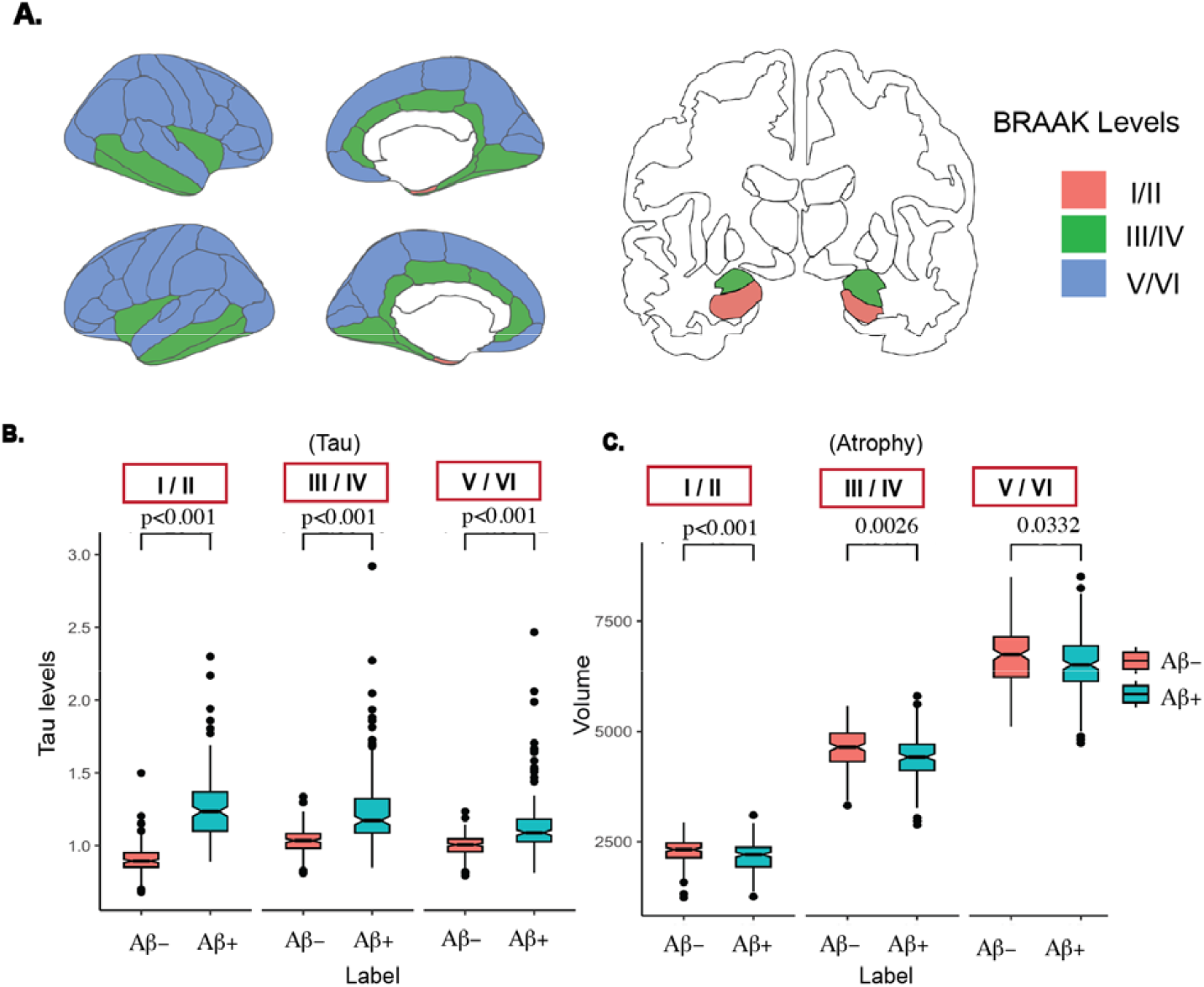
Group comparisons for tau load and atrophy (A.) Analysis was performed separately for regions of interest (ROIs) grouped according to the transentorhinal (stages I/II), limbi (stages III/IV) and isocortical (stages V/VI) Braak stages. (B.) The Aβ+ group showed greater tau uptake than the Aβ-group in ROIs corresponding to Braak I/II, Braak III/ IV and Braak V/VI. (C) The Aβ+ group also showed higher atrophy levels when compared to the Aβ -group in the same grouped ROIs.

### Braak Staging

To facilitate the interpretability of the reported results and link them with existing knowledge on the spread of pathology along the AD continuum, we compared atrophy and tau load, as well as coupling between the two, across ROIs known to be affected at different stages along Braak’s staging scheme (Braak et al., 2011, 2006; Braak and Braak, 1995) **(Supplementary Figure 1B)**. The analysis compared tau load, atrophy, and coupling between the two in ROIs grouped across the transentorhinal (stages I and II), limbic (stages III and IV), and isocortical (stages V and VI) stages (Braak and Braak, 1991).

## Statistical Analysis

Group differences in demographic data (age, gender, education) were analyzed using t-tests or chi-squared tests. These analyses were conducted in R, with the packages dgof (v1.4) (https://CRAN.R-project.org/package=dgof). Group differences in tau load and in atrophy, as well as in regional coupling between tau and atrophy were based on a non-parametric, permutation-based analysis of variance, adjusted for age (see **Table 1**). The resulting p-values were False Discovery Rate (FDR) corrected across ROIs. These tests were carried out using the aovperm function, which is part of the permuco (v1.1.1) library (https://github.com/jaromilfrossard/permuco) in R. Finally, we examined whether the association between Aβ burden and cognition, assessed with clinical dementia rating sum of boxes (CDR-SB) scores (O’Bryant, 2008), was mediated by the extent of coupling between tau and atrophy. Parallel mediation analyses were conducted in Python 3, using the pingouin package (Version 0.5.3) (https://pingouin-stats.org/). Confidence intervals in the mediation model were computed using bootstrapping (10,000 steps).

## RESULTS

### Group demographics

Our objective was to compare the extent of coupling between tau and atrophy in the presence and absence of Aβ positivity. To that effect, we divided a total of 314 individuals with MCI into two groups, Aβ + (n=158) and Aβ - (n=156), based on Aβ PET data and established cutoffs ((Landau et al., 2012b; S.M. et al., 2013), see Methods). The Aβ + and Aβ - groups did not show significant differences in sex (p=0.364) or education (p=0.536) but did differ in age (p= 0.0288) **(Table 1)**.

### Tau load and atrophy levels in Aβ + and Aβ - subjects

We first examined group difference between Aβ + and Aβ - subjects in tau load and in atrophy, grouping ROIs across the transentorhinal (stages I and II), limbic (stages III and IV), and isocortical (stages V and VI) Braak stages (Braak and Braak, 1991) **(Figure 2A)**. Age corrected mean levels of tau load **(Figure 2B)** were significantly higher in Aβ + relative to Aβ - participants in ROIs implicated in Braak stages I/II (p<0.001), III/IV (p<0.001), and V/VI (p<0.001). Similarly, age corrected mean levels of atrophy **(Figure 2C)** were significantly higher (i.e., regional volumes were lower) in Aβ + relative to Aβ - participants in ROIs corresponding to Braak stages I/II (p<0.001), III/IV (p=0.0026), and V/VI (p=0.0332). In both comparisons, significant differences were retained when outlier values were removed.

### Coupling between tau and atrophy

We next examined the regional coupling between tau and atrophy, and the extent to which it differed as a function of Aβ positivity. Taking advantage of the multilayer representation of tau and atrophy as separate layers composed of identical nodes (ROIs), we computed a coupling score between tau and atrophy, based on the Euclidean distance between the layers **(Figure 3A)**. The coupling score denoted the ratio between each spatially coupled edge and all other non-coupled edges in the multilayer network (**Figure 3B**; see Methods). In all regions corresponding to Braak I/II **(Figure 3B)**, the Aβ+ group showed significantly greater coupling (FDR corrected) in the left (p = 0.0012) and right entorhinal (p = 0.0008), as well as left (p = 0.03626) and right hippocampal (p = 0.042) ROIs. For ROIs corresponding to Braak III/IV **(Figure 3C)** the Aβ+ group showed significantly greater (FDR corrected) coupling in left lingual (p=0.017), right fusiform (p=0.009), right middle temporal (p=0.020), right insula (p=0.017) and right inferior temporal (p=0.02) ROIs. Altogether 20.8% of the regions in Braak III/IV showed significant coupling between tau and atrophy. Finally, for ROIs corresponding to Braak V/VI **(Figure 3D)** the Aβ+ group showed greater coupling (FDR corrected) in the left traverse temporal (p=0.04371), right superior frontal (p=0.008), right lateral orbitofrontal (p=0.01), right medial orbitofrontal (p=0.030), right superior parietal (p=0.028), right precuneus (p=0.008), right traverse temporal (p=0.008), right superiortemporal (p=0.008), left postcentral (p=0.03) right postcentral (p=0.008), right precentral (p=0.028), and right paracentral (p<0.008) ROIs. Overall, 27.2% of the ROIs in Braak V/VI showed significant coupling between tau and atrophy.

**Figure 3:**
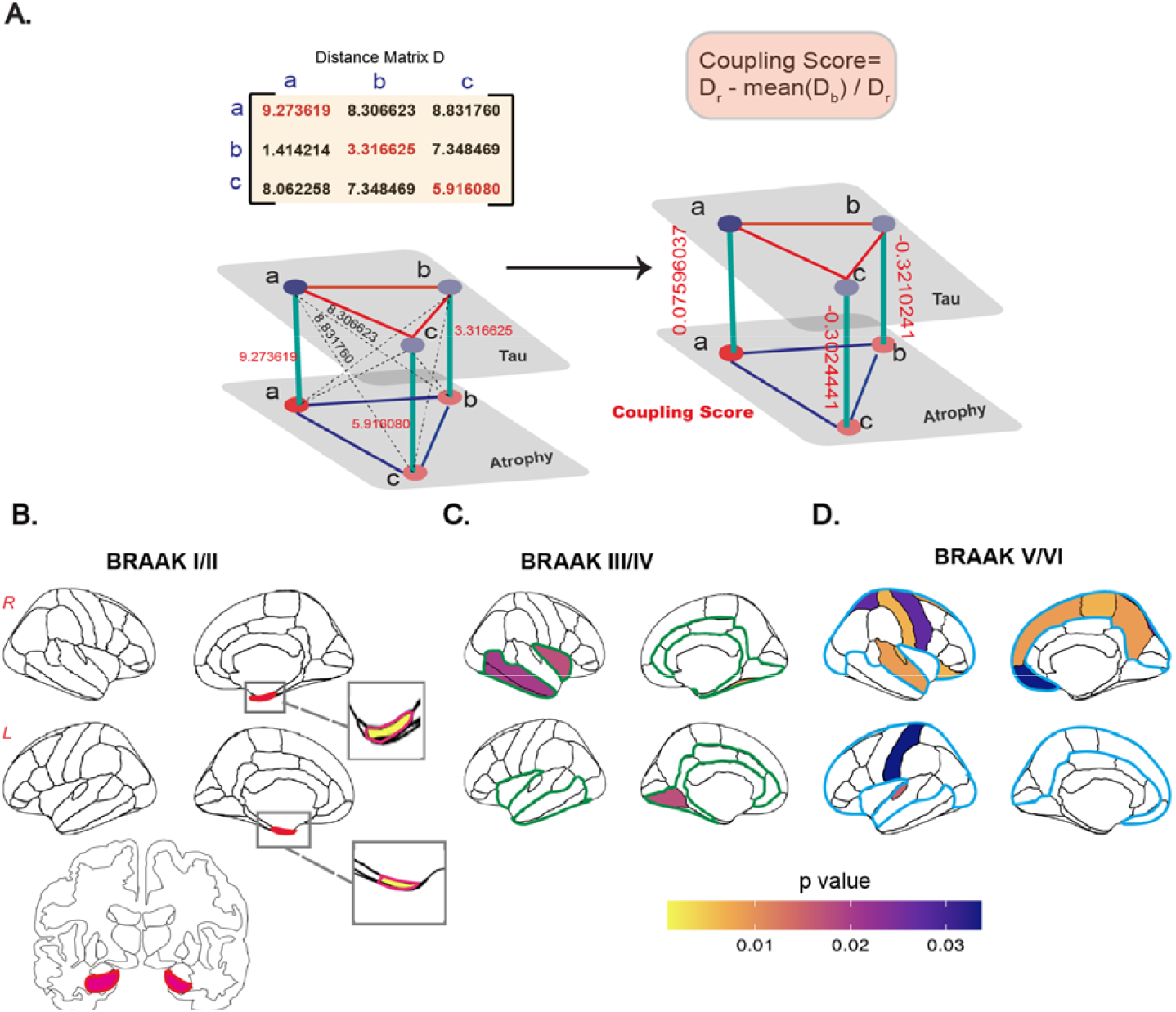
Coupling between tau and atrophy. (A.) Calculation of coupling scores computed to measure the relative coupling between tau and atrophy, was based on the Euclidean distance between layers (distance matrix D) and denotes the ratio between a spatially coupled edge D_r_ (red) and all other non-coupled edges D_b_ (black). (B.) ROIs implicated in Braak I/II stages that showed significantly higher coupling in the Aβ+ group, as compared to the Aβ-group (C.) ROIs corresponding to Braak III/IV where significantly higher coupling was found in the Aβ+ group, compared to the Aβ-group (D.) ROIs implicated in Braak V/VI that showed significantly higher coupling in the Aβ+ group, compared to the Aβ-group. Contours depict the entire set of ROIs implicated in each of the stages (corresponding to Figure 2A).

### Mediational link between Aβ burden cognition and tau-atrophy coupling

Our results so far reveal differential levels of coupling between tau and atrophy when comparing Aβ+ and Aβ-subjects. Next, we examined whether the extent of coupling and its relationship with Aβ burden also relates to subjects’ cognitive status. Considering Aβ as a continuous variable, we next tested whether the association between Aβ burden and global cognition, assessed with CDR-SB scores, were mediated by the extent of coupling between tau and atrophy (considering ROIs where coupling scores were found to be significant). This was achieved by fitting the data with a parallel mediation model. We found that coupling in the right middle temporal (p = 0.005) and right inferior temporal (p = 0.0288) ROIs (typically implicated in Braak III/IV) significantly mediated the association between of Aβ burden and CDR-SB scores **(Figure 4)**.

**Figure 4:**
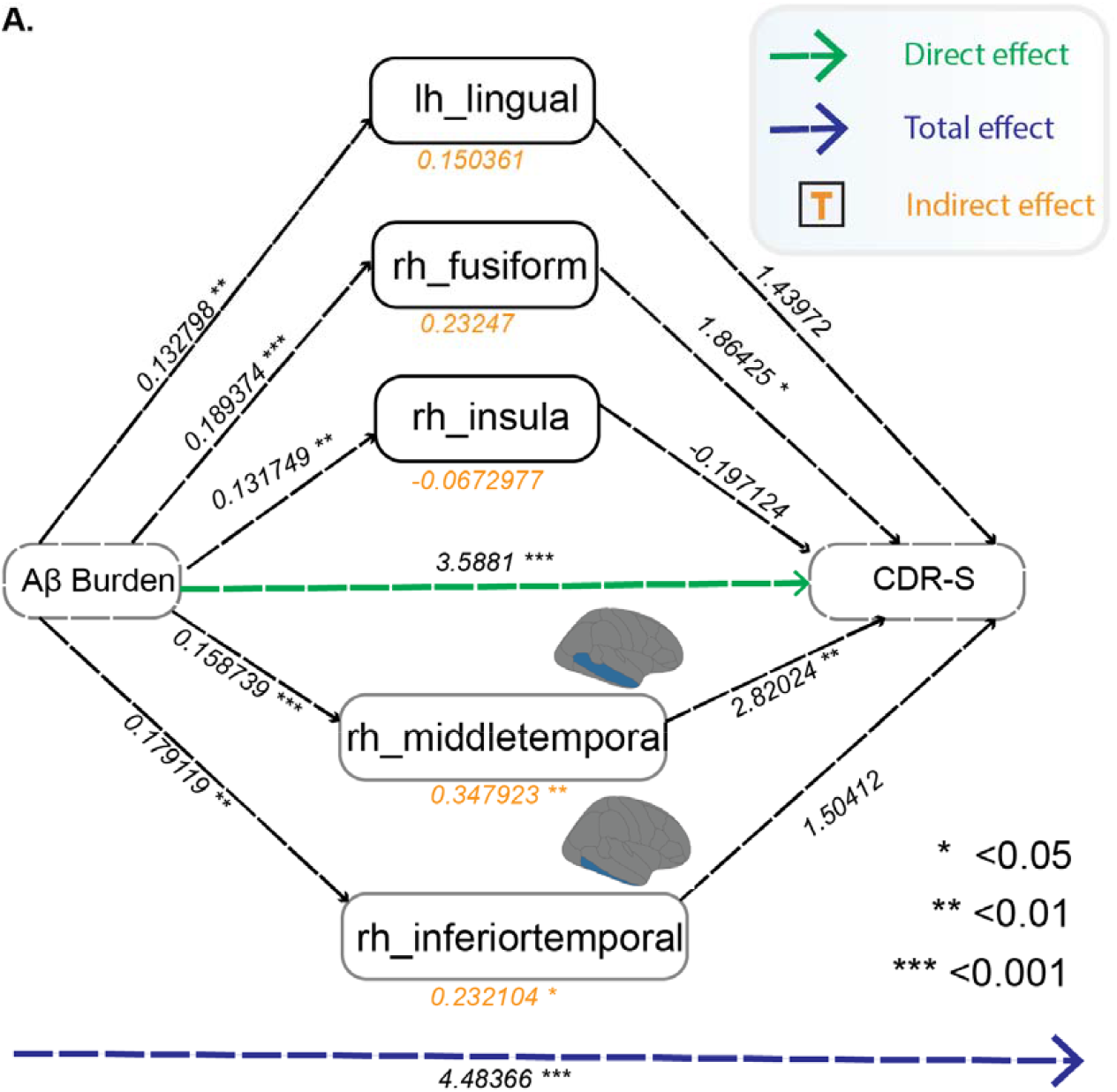
Mediational link between Aβ burden, coupling between tau and atrophy and global cognition. The association between Aβ burden, considered as a continuous variable, and global cognition (CDR-SB: Clinical Dementia Rating Scale–Sum of Boxes) was mediated by coupling between tau and atrophy in the right middle temporal (p=0.005) and right inferior temporal (p=0.02) cortices. Direct, indirect (mediation) and direct effects are shown.

## DISCUSSION

The objective of the current study was to estimate the extent of spatial coupling between tau and atrophy biomarkers in individuals with MCI, study the role of Aβ burden in this coupling, and examine the relationship between coupling and cognitive dysfunction. Overall, stronger coupling between tau and atrophy was observed in Aβ+ as compared to Aβ - individuals with MCI. The extent of coupling differences between these two groups varied spatially. Whereas strong coupling was observed in all ROIs corresponding to the transentorhinal stages (Braak stages I/II), more restricted coupling was found in ROIs from the limbic (stages III/IV) and isocortical (stages V/VI) stages. Finally, our results reveal that coupling between tau and atrophy in the middle temporal and inferior temporal cortices mediated the association between Aβ burden and cognitive dysfunction.

Stronger coupling between tau and atrophy was found in the Aβ+ group, relative to the Aβ - group in all ROIs corresponding to the early transentorhinal stages (Braak stages I/II), including the entorhinal cortex and the hippocampus. Early involvement of these regions in AD pathogenesis is well documented (Dickerson et al., 2005; Duyckaerts et al., 2015; Stoub et al., 2010). Moreover, atrophy in the hippocampus and adjacent early Braak regions is predictive of conversion from MCI to AD (Kwak et al., 2022b, 2022a). Association between tau and atrophy in regions corresponding to early Braak stages are also evident in Aβ-CN individuals (Das et al., 2019), however the current results demonstrate that coupling was stronger when Aβ load was greater. Furthermore, the strong local coupling observed here in early Braak regions is consistent with the observation that in MCI and AD, stronger local, but not distant coupling exists between tau PET load and cortical atrophy in these regions (Timmers et al., 2019).

In regions considered as being part of the limbic (stages III/IV) and isocortical (stages V/VI) Braak stages, coupling between tau and atrophy was more restricted. Altogether, approximately 25% of the ROIs affected at these mid and later Braak stages showed stronger tau-atrophy coupling in the Aβ+, relative to the Aβ - group. Of note, group differences in atrophy in ROIs corresponding to stages V/VI were rather small, thus any findings on coupling seen among these ROIs should be interpreted with caution. The more restricted level of local coupling between tau and atrophy in ROIs corresponding to later Braak stages is consistent with earlier results (Timmers et al., 2019). Considering that individuals with MCI, who are along the AD continuum, are likely at stages III/IV (Therriault et al., 2022), lower levels of coupling in ROIs corresponding to higher Braak stages is expected. Additional research is needed to further delineate the correspondence between neuropathological burden in MCI and the coupling observed between *in-vivo* imaging biomarkers.

We report that the association between Aβ burden and global cognition, as captured by CDR-SB scores is mediated by the extent of tau-atrophy coupling in right middle temporal and right inferior temporal cortices. Early involvement of these cortical regions in AD pathology has been reported (Halawa et al., 2019). Moreover, middle, and inferior temporal regions, and ROIs belonging to Braak stage III in general, were noted as critical regions in rapid conversion from MCI to AD (Morbelli et al., 2017; Woodworth et al., 2022). Altogether, our findings join these earlier observations in highlighting the contribution of pathology in the temporal lobe to cognitive dysfunction at the prodromal phases on AD.

In the current study we queried the extent of coupling between tau and atrophy by modeling multimodal neuroimaging data as a multilayer network. Multilayer networks can aid in modeling complex interactions that occur among biological (or non-biological) processes that operate at differing spatial and temporal scales (Robitaille et al., 2021). This approach may thus properly capture the heterogeneity often observed in biological systems which may result from the diverse interactions of the system’s various substrates (Hammoud and Kramer, 2020). Here, the multilayer representation allowed us to compare coupled versus non-coupled interactions among 2 distinct biological processes characteristic of the AD continuum. Future work can focus on other processes and mechanisms which can be quantified in multilayer networks, such as changes in modularity (Taylor et al., 2017; Wilson et al., 2017) redundancy (Radicchi and Bianconi, 2017), and robustness (Kumar and Singh, 2020; Liu et al., 2020), known to be strongly impacted by aging and dementia (Contreras et al., 2019; Langella et al., 2021; Sadiq et al., 2021; Song et al., 2014; Stanford et al., 2022). Moreover, studies utilizing longitudinal data will be required to better delineate changes that occur in the coupling between tau and atrophy along the AD continuum.

## Summary

In summary, we report that coupling between tau and atrophy existed throughout the brain but was more prominent in regions that are known to be affected at the transentorhinal stages of AD pathology. Altogether, these results highlight the centrality of coupling between tau and atrophy in MCI.

## Supporting information

Robinson-SM

## Data Availability

All data produced are available online at: https://adni.loni.usc.edu/

## ACKNOWLEDGMENTS AND DISCLOSURES

We thank William for valuable feedback. Research reported in this publication was supported by the National Institute on Aging of the National Institutes of Health under Award Number R01AG062590. We would also like to acknowledge NSF DMS-2113662 and NSF RTG grant DMS-2134107 for their support. The content is solely the responsibility of the authors and does not necessarily represent the official views of the National Institutes of Health. Data used in preparation of this article were obtained from the Alzheimer’s Disease Neuroimaging Initiative (ADNI) database (adni.loni.usc.edu). As such, the investigators within the ADNI contributed to the design and implementation of ADNI and/or provided data but did not participate in analysis or writing of this report. A complete listing of ADNI investigators can be found at: http://adni.loni.usc.edu/wp-content/uploads/how_to_apply/ADNI_Acknowledgement_List.pdf

