## Supplementary material for "The spatial distribution of coupling between tau and neurodegeneration in amyloid-β positive mild cognitive impairment": Robinson-SM

Eran Dayan,

Associate Professor of Radiology,

University of North Carolina at Chapel Hill.

**Short/running title.**

**Coupling between tau and neurodegeneration in MCI**

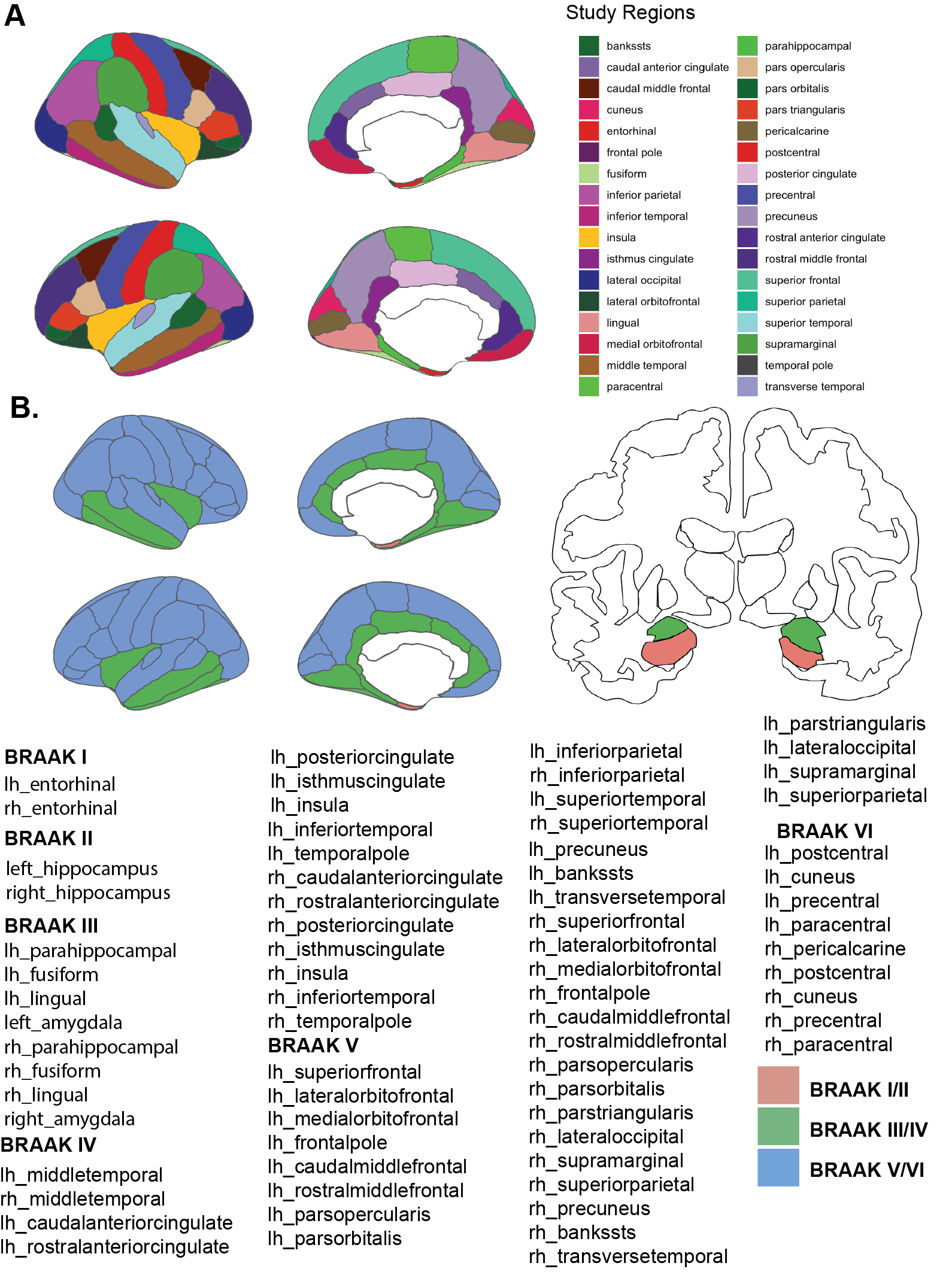

**Supplementary Figure 1** - A: The regions that are used for the study. B: Region cluster based on BRAAK levels (BRAAK I/II, BRAAK III/IV, BRAAK V/VI)

**Calculation of Coupling Score:**

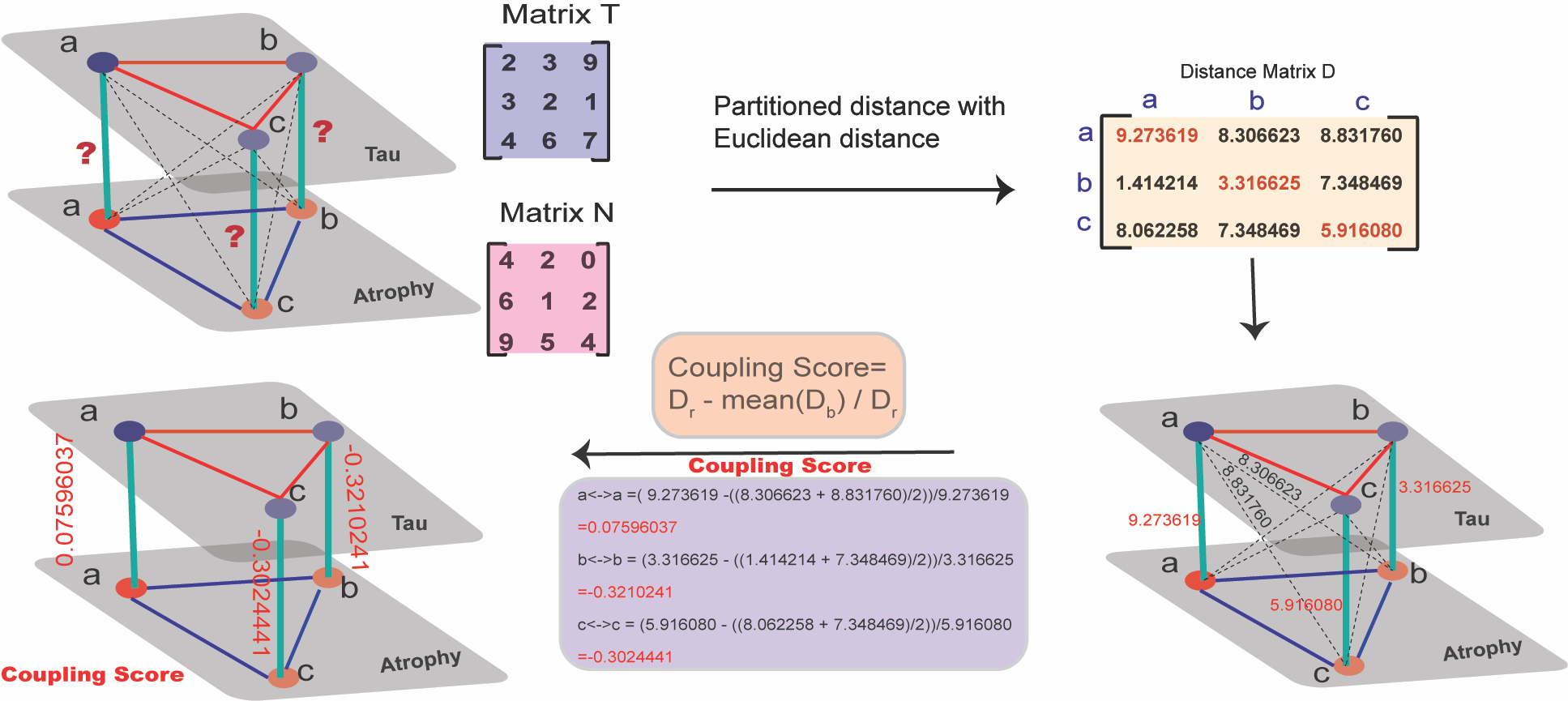

**Supplementary Figure 2:** Coupling Score calculation Toy example: The correlation matrix of tau (T) and atrophy layers (N) was subjected to partitioned distance calculation (Euclidean) to derive the distance matrix D. Distance matrix D was used to calculate coupling score.
